# Peer-led sexual and reproductive health education and risky sexual behavior among adolescent girls and young women in rural Uganda: a quasi-experimental pre-post study

**DOI:** 10.64898/2026.03.10.26348102

**Authors:** Alimah Komuhangi, Saidi Appeli, Jonathan Izudi

## Abstract

Adolescent girls and young women face significant sexual and reproductive health (SRH) challenges. We assessed the preliminary effectiveness of a community-based, peer-led SRH education intervention on risky sexual behaviors and comprehensive SRH knowledge among adolescent girls and young women aged 15-24 years in Moroto District, northeastern Uganda.

From October 2024 to January 2025, we conducted a pre-post quasi-experiment study without a comparison group across six villages. Participants were selected through multi-stage sampling and assessed at baseline. They subsequently received the community-based peer-led SRH education intervention; each participant served as her own control in the absence of a comparison group. Risky sexual behavior was the primary outcome, and comprehensive SRH knowledge was the secondary outcome. The intervention effect was assessed using a generalized estimating equation with a Poisson distribution, log link function, and exchangeable correlation structure. We reported adjusted risk ratios (aRR) with 95% confidence intervals (CI).

Of 389 participants who completed both the pre- and post-intervention evaluation, the mean ages were comparable (19.29 ± 2.94 years vs. 19.31 ± 2.91 years; p = 0.922, respectively). After the intervention, there was a significant decline in the proportion of participants who engaged in risky sexual behavior (57.1% before vs. 37.8% after, p<0.001) and a significant improvement in comprehensive SRH knowledge (85.6% before vs. 99.5% after, p<0.001). In cause-effect analysis, there was a 33% reduction in risky sexual behavior (aRR 0.67, 95% CI: 0.57-0.75), and a 16% improvement in comprehensive SRH knowledge (aRR 1.16, 95% CI: 1.12-1.20).

A community-based, peer-led SRH education intervention reduces risky sexual behavior and improves comprehensive SRH knowledge. These findings should be considered preliminary, as robust studies are needed, including a need for nuanced strategies to address contextual factors that contribute to risky behavior despite improved comprehensive SRH knowledge.

## Background

Risky sexual behavior is a significant public health challenge due to its association with negative outcomes such as sexually transmitted infections (STIs), including human immunodeficiency virus (HIV), unwanted pregnancies, and unsafe abortions [1]. Risky sexual behavior is defined as engaging in practices such as multiple sexual partners, condomless sex, sexual intercourse with commercial sex workers, or sexual intercourse under the influence of substances within a specified timeframe [2]. Addressing these behaviors is a priority for improving sexual and reproductive health (SRH) outcomes, particularly in resource-constrained settings.

Sub-Saharan Africa bears a disproportionate burden of STIs and HIV, with adolescents and young adults being the most affected [3]. According to the 2025 UNAIDS Global AIDS Update, in 2024 an estimated 210,000adolescent girls and young women (aged 15–24) acquired HIV, with approximately 3,300 of the weekly new infections occurring in sub-Saharan Africa [4]. Several factors contribute to risky sexual behavior in the region, including limited access to comprehensive sexual education, high rates of poverty, gender inequality, and sociocultural norms that may hinder the adoption of safe sexual practices [5]. Evidence from high-income countries such as the United States highlights that peer-led SRH education intervention is effective in reducing risky sexual behaviors among adolescent girls and young women [6]. However, there is limited evidence on the effectiveness of such interventions in low- and middle-income countries such as Uganda.

Uganda has implemented various policies and frameworks to address SRH challenges, including the National Sexuality Education Framework to strengthen comprehensive sexuality education in schools [7], the Adolescent Health Policy Guidelines and Service Standards to improve adolescent-friendly SRH services [8], and the National Policy Guidelines and Service Standards for Sexual and Reproductive Health and Rights to guide national SRH service delivery [9]. Despite the implementation of these policies, SRH outcomes in certain regions, such as northeastern Uganda, remain poor: high rates of early marriages, adolescent pregnancies, and STIs. These challenges underscore the need for tailored and context-relevant interventions to address them. Community-based and peer-led intervention has shown promise in improving SRH outcomes among adolescent girls and young women in developed countries [10]. However, there is a lack of evidence about its effectiveness in resource-limited settings such as Moroto District in northern Uganda. In Moroto district, adolescent girls and young women face unique challenges that exacerbate risky sexual behaviors. The challenges include high poverty rates (40%), low literacy levels (55%), and limited access to formal education and employment opportunities [11]. Moreover, cultural norms that prioritize large families and early marriage further increase the vulnerability of adolescent girls and young women to STIs and HIV in the setting.

To generate preliminary evidence to inform future large-scale studies, we evaluated the effectiveness of a community-based, peer-led SRH education intervention in reducing risky sexual behaviors and improving comprehensive SRH knowledge among adolescent girls and young women in rural northeastern Uganda. The intervention was conceptually informed by the socio-ecological model, which posits that improvements in SRH knowledge and peer support can influence individual sexual health behaviors [15].

## Materials and methods

### Study setting

The study was conducted across seven villages in Moroto district in northeastern Uganda. The seven villages included Bazar, Camp Swahili, Kakoliye, Labour Line, Nakapelimen, Natumukasko, and Singila. Moroto district has limited educational infrastructure, with only 12 secondary schools and 45 primary schools serving a population of over 200,000 [12]. The high student-teacher ratio (45:1) and high dropout rates (30%) further limit access to SRH education, leaving many young people without adequate knowledge of safe sexual practices.

### Study design and population

We employed a pre-post quasi-experimental study without a comparison group to examine the effectiveness of a community-based, peer-led SRH education intervention in reducing risky sexual behaviors and improving SRH knowledge. The design was ideal for this pilot study, as it allowed for an initial assessment of the intervention’s effectiveness without the added complexity of a comparison group [13]. The study population consisted of adolescent girls and young women aged 15-24 years. Participants were eligible if they had lived in the area for at least six months. To strengthen causal inference, the same individuals were included in the pre-and-post evaluation. This means that we had a repeated measures (longitudinal) design, where each participant served as their own control. We excluded adolescent girls and young women who had visited the areas for a stay lasting less than six months, including those with known mental disorders (based on medical records verification), as they would have difficulty understanding the intervention.

### Sampling approach

This study utilized a multi-stage sampling technique to select the participants. First, the district was purposely selected, and six villages within the district were selected using a simple random sampling approach based on the list of all villages. Next, the local village-level records were used to obtain a sampling frame for households with adolescent girls and young women. Here, a list of all eligible households was prepared, and each was assigned a unique number. Using a computer-generated random number table, participants were randomly selected to achieve the required sample size. In households with more than one eligible adolescent girl, one participant was randomly selected using a simple lottery method.

### Data collection

We collected data at the baseline and endline. The baseline and endline data were collected in October 2024 and January 2025, respectively, by administering a researcher-administered questionnaire focused on the following: 1) characteristics of participants, 2) risky sexual behavior, and 3) SRH knowledge. The questionnaire was developed to suit the context (Supplementary file 1).

### Intervention

The intervention was a community-based, peer-led SRH education package implemented in the six selected villages. The intervention content drew on the World Health Organization (WHO) (2018) Guidelines on Adolescent SRH, emphasizing risk prevention, empowerment, and communication competence [14]. The intervention delivery was informed by preceding qualitative research on barriers and facilitators to implementation [15]. The intervention comprised four interactive modules delivered over three months, from 12 October 2024 to 12 January 2025 (Table 1). Two modules were delivered during the first month through a combined extended session, with a deliberate two-week interval between the delivery of the first and second modules to allow for reflection and consolidation of learning. The remaining two modules were delivered one per month during the second and third months. This sequencing allowed full coverage of all four modules within the planned three-month intervention period while maintaining manageable session lengths and participant engagement. Each session lasted approximately 60–90 minutes, and all sessions were conducted in accessible community venues, including community halls and youth centres.

**Table 1:** Summary of the intervention modules and delivery approaches.

| Module | Title/focus | Timing | Content/activities | Delivery method/notes |
| --- | --- | --- | --- | --- |
| 1 | Prevention of risky sexual behaviour | Month 1 | Introduced concepts of bodily autonomy, consent, sexual decision-making, and risky behaviour. Activities included role-plays and peer dialogues using locally salient scenarios of early marriage and transactional sex identified in the formative phase. | Peer-led sessions; bilingual (English and Ngakarimojong) |
| 2 | Prevention of unwanted pregnancies, unsafe abortions, and HIV/STI transmission | Month 1 | Integrated factual biomedical knowledge with culturally adapted myths-debunking sessions to address prevalent fertility myths (“contraceptives burn eggs”) and stigma. Visual aids and storytelling were used to correct misinformation and build normative support for contraceptive uptake. | Peer-led sessions; bilingual (English and Ngakarimojong) |
| 3 | Gender-based violence and communication skills | Month 2 | Focused on interpersonal negotiation, refusal skills, and conflict resolution using WHO's life-skills framework. Discussions linked gender norms, power imbalances, and violence prevention to broader notions of respect and mutual responsibility drawn from local idioms. | Peer-led sessions; bilingual |
| 4 | Contraceptive methods and sexual-health counselling | Month 3 | Demonstrated modern contraceptive options and referral processes to youth-friendly services. Sessions simulated clinical interactions to build confidence and reduce fear of judgmental providers. | Peer-led sessions; bilingual; participants received nominal transport reimbursement |

Participants were mobilised through peer-led community outreach and existing social networks, including referrals by peer educators, community meetings, and word-of-mouth within adolescent girls and young women’s social groups, with support from local community leaders and youth focal persons. The intervention was delivered by six trained female peer educators aged 18–24 years from the same communities as participants, all of whom had at least secondary-level education and prior experience in sexual and reproductive health (SRH) peer education. Peer educators completed a three-day training of trainers facilitated by public health professionals with expertise in SRH, adolescent health, and community-based interventions. The training of trainers covered intervention content, facilitation skills, ethical and safeguarding considerations, referral pathways, and participatory learning methods. Ongoing supervision and mentorship were provided to ensure consistency of delivery. To enhance participation and adherence, reminder follow-ups were conducted through peer networks, and light refreshments were provided during sessions to encourage consistent attendance and engagement. No adverse events or unintended negative effects related to the intervention were reported during the study period.

### Study outcomes

The primary outcome was risky sexual behavior measured on a binary scale (yes or no), measured using: 1) multiple sexual partners measured using data on the number of sexual partners in the last three months, categorized as < 2 or ≥ 2; 2) condomless sex in the past three months determined using data on whether or not the participant had used a condom at all sexual encounters, categorized as yes or no; 3) sexual intercourse with a commercial sexual sex worker in the past three months (yes or no), using data on whether or not the participant reported having had sex with a commercial sex worker; 4) sexual intercourse under influence of alcohol or substance abuse in the past three months (yes or no), using data on history of sex under influence of alcohol or substance abuse. An individual who provided an affirmative response to any one of the four behaviors was considered to have engaged in risky sexual behaviour, and those that do not provide an affirmative response to any of the four behaviors were considered not to have engaged in risky sexual behavior [2].

The secondary outcome was SRH knowledge, also measured on a binary scale. Knowledge of sexual and reproductive health (SRH) services among adolescent girls and young women was assessed using five indicators adapted from previous studies and the Uganda Demographic and Health Survey [12]. These indicators included awareness of at least one modern contraceptive method, which was used to measure contraceptive knowledge; having ever heard about any infection that can be acquired through sexual intercourse, which assessed knowledge of STIs; and comprehensive knowledge of HIV/AIDS, which was measured as a composite indicator comprising awareness of HIV/AIDS, knowledge of its modes of transmission, and the ability to correctly reject at least two out of four common misconceptions. The misconceptions included the belief that a healthy-looking person cannot have HIV/AIDS, HIV can be transmitted by mosquito bites, HIV/AIDS can be transmitted through supernatural means, and a person can become infected by sharing food with someone who has HIV/AIDS [12]. In addition, participants’ self-reported perceived risk of being infected with STIs, including HIV/AIDS, was assessed and categorized as “at risk” versus “not at risk,” and their perceived risk of becoming pregnant was similarly rated. Participants who reported awareness of at least one modern contraceptive method, had heard of infections transmitted through sexual intercourse, and demonstrated awareness of HIV/AIDS and its transmission while correctly rejecting at least two of the four common misconceptions were classified as having comprehensive knowledge of SRH services [12].

The outcome measures were adapted from previously published studies [1,2] and standardized population-based instruments, including the Uganda Demographic and Health Survey (UDHS), which has established validity and reliability in measuring sexual and reproductive health indicators in similar populations. The UDHS HIV knowledge items and SRH indicators have demonstrated acceptable internal consistency and construct validity in sub-Saharan African settings. Before data collection, the questionnaire was reviewed by public health experts for content validity and pre-tested in a similar community to ensure clarity, contextual appropriateness, and cultural relevance. Necessary adjustments were made to improve comprehension without altering the conceptual meaning of the items.

### Covariates

The covariates included sociodemographic characteristics (age, religion, marital status, age at marriage, schooling status, employment status, and highest level of education attained), family characteristics (parental survival status), and sexual and reproductive health–related characteristics (ever had sexual intercourse, age at sexual debut, unmet need for family planning, and self-reported history of sexually transmitted infection in the past six months). These variables were included in multivariable generalized estimating equation (GEE) models to adjust for potential confounding and to estimate the independent effect of the intervention on risky sexual behavior and comprehensive sexual and reproductive health knowledge.

### Statistical methods

We hypothesized that the intervention would reduce risky sexual behavior. Therefore, the sample size was calculated based on McNemar’s test of paired proportions using an online calculator, with the outcome as risky sexual behavior. The probability of discordant pairs before and after the intervention was taken as 30% vs. 20%, respectively. Accordingly, at 80% statistical power, the estimated sample size was 389 pairs assuming a 5% statistical significance level. We summarised numerical data as means and standard deviations (SD) when approximately normally distributed, or as medians and interquartile ranges (IQR) when skewed. Categorical data were summarized as frequencies and percentages. For paired pre-post comparisons, we applied the paired t-test for numeric data when the differences between pre- and post-measurements were approximately normally distributed, and the Wilcoxon signed-rank test when this assumption was violated. For categorical binary outcomes, we used McNemar’s test to compare paired proportions, while for categorical outcomes with more than two levels, we employed the Stuart-Maxwell test for marginal homogeneity. To assess the effect of the intervention on the outcomes while adjusting for baseline covariates, we applied a GEE with a Poisson distribution, log link function, and exchangeable correlation structure. We reported adjusted risk ratios (aRR) with their 95% confidence intervals (CI). All statistical analyses were conducted using Stata version 17.0 (StataCorp LLC, College Station, TX, USA). All 389 participants enrolled at baseline were successfully followed up and reassessed at endline, resulting in complete paired data for analysis. Consequently, no imputation of missing data was required. All participants were included in the final analysis, and the analyses were conducted on the full cohort consistent with an intention-to-treat approach.

### Ethical consideration

The study received ethical approval from Clarke International University Research Ethics Committee (CLARKE-2024-1113) and the Uganda National Council for Science and Technology (HS4697ES). Written informed consent was obtained from participants. For minors aged 15–17 years, assent was obtained from the participant and consent from their parent or guardian in accordance with local ethics requirements.

## Results

### Distribution of participants’ characteristics before and after the intervention

Table 2 summarizes the participants’ characteristics before and after the intervention. The mean age of participants was comparable both before and after the intervention (19.29 ± 2.94 years vs. 19.31 ± 2.91 years; p = 0.922), and the same applies to the mean age between those ever married (6.33 ± 2.22 years vs. 6.51 ± 2.30 years; p = 0.465). About one-quarter of participants were in school (24.7%), and roughly 19% were employed at both time points. Most participants had primary or secondary education, while a few had tertiary or university-level education. Parental status was similar, with most participants reporting both parents alive (approx. 58%). About 73% of the participants had sexual intercourse, with similar mean age at first sexual intercourse before and after intervention. The majority of the pregnancies were unplanned (approximately 79–80%). Unmet need for family planning was reported by roughly 12% of participants, and about 10% reported a sexually transmitted infection in the past six months. No statistically significant differences were observed in any participant characteristics before and after the intervention (all p > 0.05).

**Table 2:** Distribution of participants’ characteristics before and after the intervention.

| Variables | Level | Before intervention (n=389) | After intervention (n=389) | P-value |
| --- | --- | --- | --- | --- |
| Age (years) | Mean (SD) | 19.29 (2.94) | 19.31 (2.91) | 0.922 |
| Religion | Christian | 351 (90.2) | 351 (90.2) | 1.000 |
|  | Muslim | 35 (9.0) | 35 (9.0) |  |
|  | Other religion | 3 (0.8) | 3 (0.8) |  |
| Marital status | Widowed | 2 (0.5) | 2 (0.5) | 0.786 |
|  | Separated/divorced | 17 (4.4) | 15 (3.9) |  |
|  | Single | 210 (54.0) | 198 (50.9) |  |
|  | Married/cohabiting | 160 (41.1) | 174 (44.7) |  |
| Age at marriage | Mean (SD) | 6.33 (2.22) | 6.51 (2.30) | 0.465 |
| Type of employment | Employed | 74 (19.0) | 72 (18.5) | 0.927 |
|  | Not employed | 315 (81.0) | 317 (81.5) |  |
| In-school | No | 292 (75.3) | 292 (75.3) | 1.000 |
|  | Yes | 96 (24.7) | 96 (24.7) |  |
| Highest level of education | None | 36 (9.3) | 42 (10.8) | 0.902 |
|  | Primary | 196 (50.4) | 192 (49.4) |  |
|  | Secondary | 130 (33.4) | 132 (33.9) |  |
|  | Tertiary institution | 17 (4.4) | 13 (3.3) |  |
|  | University | 10 (2.6) | 10 (2.6) |  |
| Parental status | Only mother alive | 91 (23.4) | 87 (22.4) | 0.952 |
|  | Only father alive | 22 (5.7) | 24 (6.2) |  |
|  | Both father and mother are alive | 226 (58.1) | 224 (57.6) |  |
|  | Both Father and mother are dead | 50 (12.9) | 54 (13.9) |  |
| Ever had sexual intercourse | No | 104 (26.7) | 100 (25.7) | 0.807 |
|  | Yes | 285 (73.3) | 289 (74.3) |  |
| Age at first sexual intercourse among ever had sex | Mean (SD) | 16.36 (1.97) | 16.30 (2.03) | 0.717 |
| Type of pregnancy | Planned | 36 (20.5) | 40 (22.0) | 0.823 |
|  | Not planned | 140 (79.5) | 142 (78.0) |  |
| Unmet need for FP | No | 340 (87.9) | 342 (88.4) | 0.912 |
|  | Yes | 47 (12.1) | 45 (11.6) |  |
| Had an STI in the past 6 months | No | 348 (89.9) | 348 (89.9) | 1.000 |
|  | Yes | 39 (10.1) | 39 (10.1) |  |

### Sexual risk profiles and risky sexual behavior among participants before and after the intervention

Table 3 presents the distribution of sexual risk profiles and risky sexual behavior among adolescent girls and young women before and after the intervention. Overall, there was a substantial reduction in risky sexual behavior following the intervention. The proportion of adolescent girls and young women with multiple sexual partners in the past three months significantly decreased from 32.6% before to 14.4% after the intervention (p<0.001). Similarly, the proportion reporting condomless sex in the past three months declined from 47.8% before to 37.5% after (p = 0.005). There were no differences in the proportions of adolescent girls and young women who reported having sex with a commercial sex worker (1.0% both before and after) or having sex while intoxicated (9.8% both before and after; p = 1.000 for both). The overall risky sexual behavior significantly decreased from 57.1% before to 37.8% after the intervention (p < 0.001).

**Table 3:**
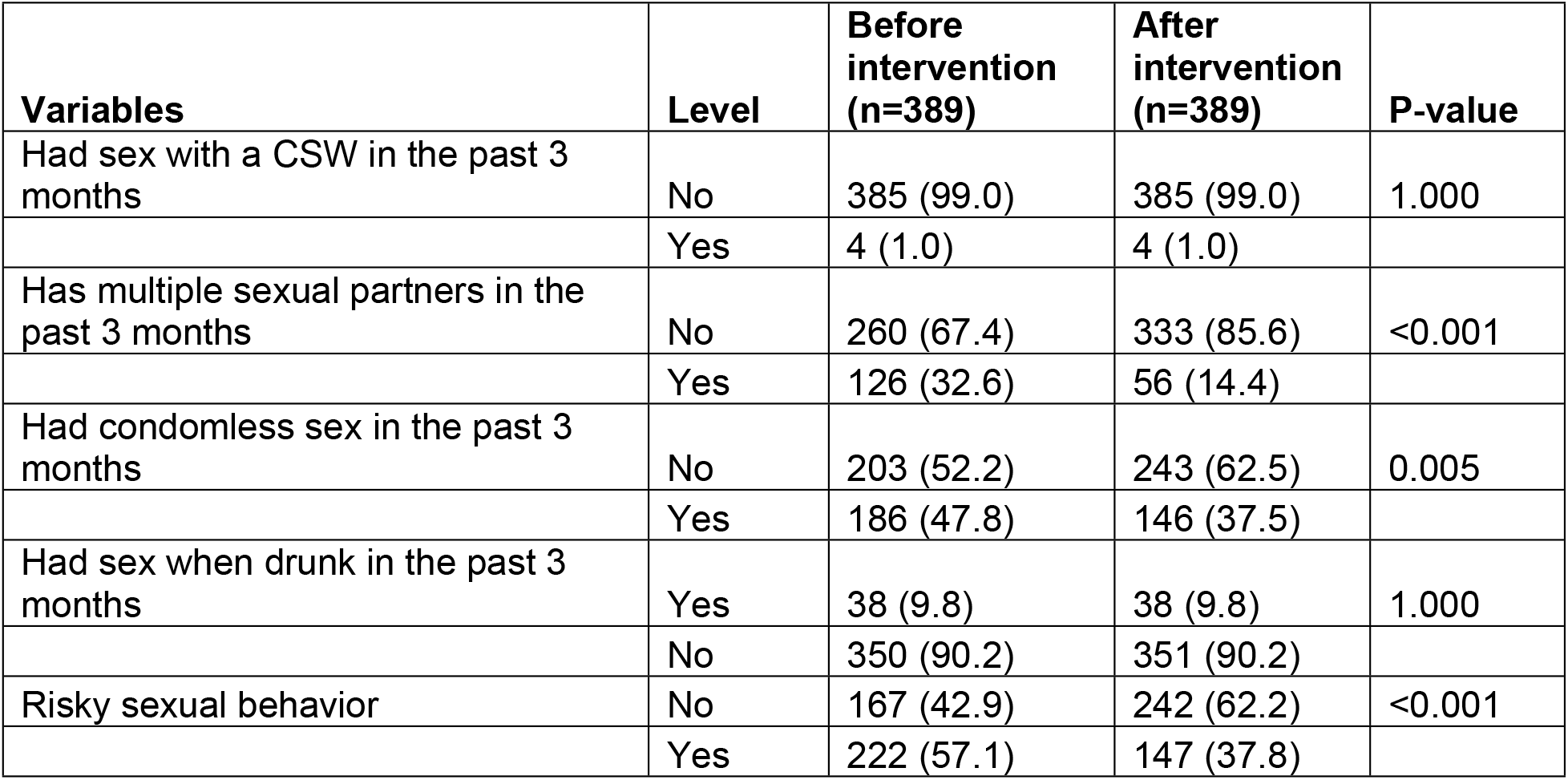
Sexual risk profiles and risky sexual behavior among participants before and after the intervention.

| Variables | Level | Before intervention (n=389) | After intervention (n=389) | P-value |
| --- | --- | --- | --- | --- |
| Had sex with a CSW in the past 3 months | No | 385 (99.0) | 385 (99.0) | 1.000 |
|  | Yes | 4 (1.0) | 4 (1.0) |  |
| Has multiple sexual partners in the past 3 months | No | 260 (67.4) | 333 (85.6) | <0.001 |
|  | Yes | 126 (32.6) | 56 (14.4) |  |
| Had condomless sex in the past 3 months | No | 203 (52.2) | 243 (62.5) | 0.005 |
|  | Yes | 186 (47.8) | 146 (37.5) |  |
| Had sex when drunk in the past 3 months | Yes | 38 (9.8) | 38 (9.8) | 1.000 |
|  | No | 350 (90.2) | 351 (90.2) |  |
| Risky sexual behavior | No | 167 (42.9) | 242 (62.2) | <0.001 |
|  | Yes | 222 (57.1) | 147 (37.8) |  |

### Comprehensive SRH knowledge among participants before and after the intervention

Table 4 presents the distribution of SRH knowledge indicators among adolescent girls and young women before and after the intervention. Overall, the level of accurate SRH knowledge improved markedly after implementation. Misconceptions about HIV transmission decreased substantially. The proportion of adolescent girls and young women who believed that HIV can be transmitted by supernatural means declined from 15.3% before to 7.8% after the intervention (p < 0.001). Misconceptions about HIV transmission through sharing food declined significantly following the intervention. The proportion of participants correctly reporting that HIV cannot be transmitted through sharing food increased from 7.4% at baseline to 11.9% post-intervention, indicating a significant improvement in understanding (p < 0.001). Knowledge that HIV cannot be transmitted through mosquito bites was already high at baseline (96.7%) and showed only a small, non-significant increase after the intervention (98.2%; p = 0.292). Similarly, awareness that HIV can be transmitted through condomless sexual intercourse with a person living with HIV remained high before (88.4%) and after (92.5%) the intervention, with no statistically significant change (p = 0.069). In contrast, knowledge related to pregnancy risk improved markedly. The proportion of adolescent girls and young women who correctly understood that pregnancy can occur following a single act of unprotected sexual intercourse increased from 85.8% at baseline to 97.4% post-intervention (p < 0.001). Overall, comprehensive sexual and reproductive health knowledge increased significantly from 85.6% before the intervention to 99.5% after the intervention (p < 0.001).

**Table 4:** Comprehensive SRH knowledge among participants before and after the intervention.

| Variables | Level | Before intervention (n=389) | After intervention (n=389) | P-value |
| --- | --- | --- | --- | --- |
| HIV is transmissible by mosquitoes | No | 354 (96.7) | 380 (98.2) | 0.292 |
|  | Yes | 12 (3.3) | 7 (1.8) |  |
| HIV is transmissible by supernatural means | No | 310 (84.7) | 30 (7.8) | <0.001 |
|  | Yes | 56 (15.3) | 356 (92.2) |  |
| HIV is transmissible through the sharing of food | No | 338 (92.6) | 46 (11.9) | <0.001 |
|  | Yes | 27 (7.4) | 340 (88.1) |  |
| HIV is transmissible through condomless sex with a person with HIV | No | 45 (11.6) | 29 (7.5) | 0.069 |
|  | Yes | 344 (88.4) | 359 (92.5) |  |
| You can get pregnant with one act of sexual intercourse without contraception. | No | 55 (14.2) | 10 (2.6) | <0.001 |
|  | Yes | 331 (85.8) | 379 (97.4) |  |
| Comprehensive SRH knowledge | No | 56 (14.4) | 2 (0.5) | <0.001 |
|  | Yes | 333 (85.6) | 387 (99.5) |  |

### Effect of the intervention on risky sexual behavior and comprehensive SRH knowledge among the participants

At endline, the proportion of adolescent girls and young women engaging in risky sexual behavior declined markedly from 57.1% before the intervention to 37.8% after. In the multivariable GEE model, the intervention was associated with a 33% reduction in risky sexual behavior (adjusted risk ratio (aRR) 0.67, 95% CI: 0.57-0.75). Conversely, the proportion of adolescent girls and young women with adequate comprehensive SRH knowledge increased substantially, from 85.6% before to 99.5% after the intervention. The intervention improved SRH knowledge by 16% (aRR 1.16, 95% CI: 1.12-1.20) as indicated in Table 5.

**Table 5:**
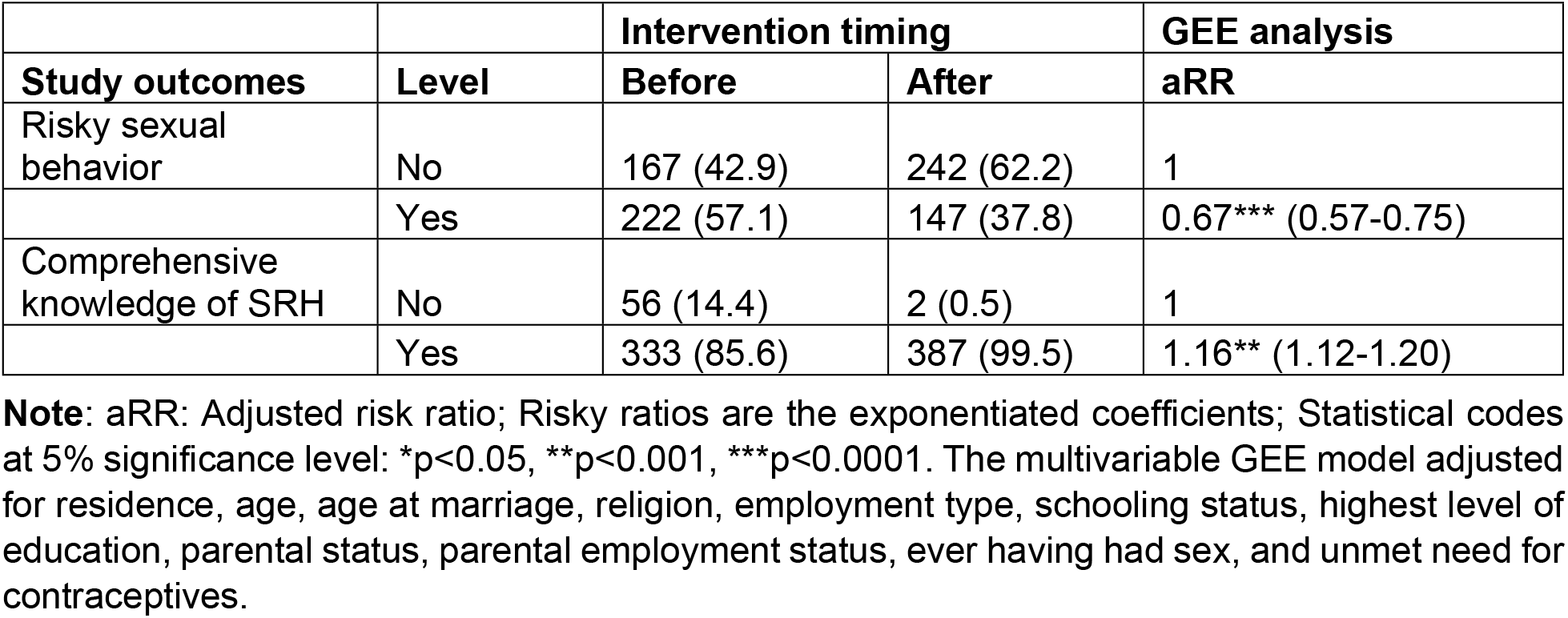
Effect of intervention on risky sexual behavior and comprehensive SRH knowledge among the participants.

## Discussion

In this study, we evaluated the effectiveness of a community-based peer-led sexual and reproductive health education intervention in reducing risky sexual behaviors and improving comprehensive SRH knowledge among adolescent girls and young women aged 15–24 years in rural northeastern Uganda. We found a significant reduction in risky sexual behavior and an improvement in comprehensive SRH knowledge among adolescent girls and young women. The decrease in risky sexual behavior in this study aligns with previous literature that highlights the effectiveness of targeted interventions aimed at promoting safer sexual practices. For instance, studies conducted in low- and middle-income countries show that educational interventions influence behavior change, particularly among adolescents and young adults, by improving their understanding of risks associated with unsafe sexual practices [16;17;18].

Relatedly, in northern Uganda, socio-cultural and peer-led education approaches have been emphasized in reducing risky sexual behaviors [19, 2]. Also, among adolescent girls and young women, community-based interventions that integrate sexual and reproductive health education with supportive social environments have been shown to enhance the adoption of positive behaviors such as consistent condom use, delayed sexual debut, reduction in multiple sexual relations, and increased utilization of sexual and reproductive health services [20]. Such interventions have also been associated with reduced alcohol consumption [21], which is a key driver of risky sexual behavior. Additionally, in a systematic review, school-based and community-driven sex education interventions demonstrated effectiveness in preventing HIV infection in low- and middle-income countries [18]. The significant reduction in risky sexual behavior in this study, therefore, supports the argument that targeted educational programs play a pivotal role in addressing sexual health risks and behavior among adolescent girls and young women.

The finding that the intervention significantly improved comprehensive SRH knowledge among adolescent girls and young women is consistent with evidence from systematic reviews indicating that peer-led education is an effective strategy for improving adolescents’ knowledge of SRH topics [22]. Empirical evidence from similar contexts further supports these findings. For instance, a peer-led SRH education program implemented in Rwanda reported a significant increase in SRH knowledge among students, with post-intervention test scores substantially higher than baseline levels [23]. Similarly, a recent cluster-randomized trial in Zambia (the Yathu Yathu study) demonstrated that delivering SRH services through peer-led community “hubs” significantly increased the level of awareness of HIV serostatus among adolescents [24]. Collectively, these prior study findings reinforce our findings and illustrate that peer-led approaches have the potential to successfully enhance SRH knowledge across diverse sub-Saharan African settings.

Importantly, our results demonstrate that even in a remote, resource-limited region such as Karamoja, a well-designed peer-led education intervention can meaningfully improve the understanding of sexual and reproductive health among adolescent girls and young women. These findings contribute to supporting the long-standing recommendations advocating comprehensive sexuality education that actively involves communities in challenging harmful social norms [25]. They also underscore the importance of flexible and context-responsive intervention designs when addressing the diverse needs of adolescents, particularly those living in marginalized settings [26]. Consistent with findings from other peer-led interventions, the community-based peer-led SRH education sessions also helped dispel persistent myths and misconceptions surrounding fertility and contraceptive methods [27]. The improvement in comprehensive SRH knowledge, therefore, represents a meaningful increase in health literacy among a highly vulnerable population of girls who previously had limited access to accurate and youth-friendly SRH information. This enhancement in knowledge is a critical precursor to informed decision-making and equitable access to sexual and reproductive health services.

The success of the intervention may also be attributed to the community-based and peer-led delivery approach in the local language, which enhanced acceptability, cultural relevance, and engagement among participants [25].

### Study strengths and limitations

Our study has several strengths and some limitations. First, to the best of our knowledge, this is the first study in Moroto district to examine the effectiveness of a community-based peer-led sexual and reproductive health intervention in reducing risky sexual behavior and improving SRH knowledge among adolescent girls and young women. Our study acknowledges some limitations. The study relied on self-reported data regarding knowledge and risky sexual profiles, which may be subject to self-reporting bias. Additionally, the study did not include a comparison group, which limited the ability to account for time-varying confounders to strengthen causal inference. Lastly, we do not know if the improvements may extend to the post-intervention period, as we do not have the data. Future studies should, therefore, examine whether the outcomes are sustained beyond the intervention period.

## Conclusion and recommendation

This study, conducted among adolescent girls and young women aged 15–24 years in Moroto District in northeastern Uganda, found that a community-based peer-led sexual and reproductive health education intervention significantly reduced risky sexual behavior and improved comprehensive SRH knowledge. This underscores the transformative potential of peer-led, community-based approaches in resource-limited settings. Future studies are needed on nuanced strategies to address contextual factors contributing to risky behavior despite improvement in knowledge.

## Data Availability

Data will be provided upon request.

## Acknowledgements

We thank Clarke International Research Ethics Committee (CIUREC) and the Uganda National Council for Science and Technology (UNCST) for ethical approval, and the District Health Officer of Moroto District for granting us administrative clearance to conduct the study. Finally, we thank the adolescent girls and young women in Moroto District who consented to participate in this study.

## Availability of data and materials

The datasets and materials used during the current study are available upon request.

## Contributions

AK and JI: Study conception and design, AK: Acquisition of data. Analysis and interpretation of data. AK, SA, and JI: Drafting of manuscript. AK, SA and JI Critical revision. AK, SA, and JI: Final approval of manuscript.

